# Developmental brain age gap in prematurity and postnatally emerging delay in congenital heart disease

**DOI:** 10.64898/2026.04.01.26349523

**Authors:** Misha Kaandorp, Kelly Payette, Anna Speckert, Celine Steger, Hui Ji, Hosna Asma Ull, Ruth Tuura, Cornelia Hagmann, Walter Knirsch, Beatrice Latal, Jing-Ya Ren, Su-Zhen Dong, Hyun Gi Kim, Andras Jakab

## Abstract

Brain development follows a precisely regulated biological timetable, with defined periods of vulnerability increasingly recognized in congenital disorders affecting early brain development. This biological timing can be captured by the emerging concept of brain age, a measure of brain maturation, enabling the detection of deviation from normative developmental trajectories. Clinical conditions affect the degree of brain development during this critical period, including preterm birth and congenital heart disease (CHD).

We developed a deep learning-based brain age estimation framework across the fetal–neonatal period (21-44 gestational weeks) to quantify neurodevelopment from structural MRI. Using 1056 scans from six datasets acquired at three centers, Zurich, Shanghai, and the Developing Human Connectome Project, we trained models on normative fetal and neonatal MRI data. Both structural MRI-based and segmentation-derived cortical morphology-based models were implemented to assess representation effects and cross-center generalisability. The framework was applied to two clinically relevant conditions, preterm birth and CHD, to estimate the brain age gap (BAG), defined as the difference between predicted brain age and chronological age.

In preterm neonates scanned at term-equivalent age (n=90, 37-44 weeks), BAG was progressively more negative with lower gestational age at birth. Neonates born before 28 weeks showed delays of −0.7 to −0.8 weeks relative to term-born controls. In CHD (n=50, 22-34 weeks), fetal brain age did not differ from center-matched controls and no association with cardiac defect severity was observed. After birth, neonates with CHD (n=110, 37-44 weeks) showed significant (p<0.05) negative BAGs before surgery (-1.3 to -1.8 weeks) and BAGs increased significantly (p<0.05) after surgery (up to -3 weeks in center-specific analyses), indicating a delay in brain maturation from postnatal stage, but not in prenatal stage in CHD patients. These patterns were found across both structural MRI-based models and cortical morphology-based models, despite the need for cross-center calibration to minimize systematic bias. Voxel-based morphometry showed that a larger BAG was associated with regional contraction in deep frontal and peri-Rolandic white matter in preterm neonates, and perioperative spatial shifts in neonates with CHD. Saliency maps converged on deep white matter and periventricular regions, highlighting a potential link between BAG and delayed maturation of rapidly developing projection pathways.

These findings may indicate neurodevelopmental delays in preterm birth and a postnatally emerging maturational gap in CHD that increases following cardiac intervention. Despite limited generalisability of our methods, these results support a continuous fetal-neonatal brain age metric as a sensitive marker of global neurological maturational timing.

## Introduction

Human brain development across late gestation and neonatal life involves rapid changes in cortical folding, volumetric growth of neuroanatomical structures, white-matter maturation, and synaptic organisation, reflecting the sequential processes that establish the core neuroanatomical structures during this period^1^. This precisely regulated process creates vulnerability^2–4^. Congenital disorders such as congenital heart disease (CHD) and preterm birth are characterised by clinical and neurodevelopmental heterogeneity, the origins of which to be understood. In CHD, many fetuses and neonates demonstrate no overt structural abnormalities on routine brain magnetic resonance imaging (MRI), but many later develop mild to moderate cognitive and behavioural impairments^5,6^. In some conditions, outcomes cannot be attributed to focal brain injury or gross anatomical defects^7^, suggesting that more subtle alterations in global maturational timing may underlie the observed variability in neurodevelopmental trajectories.

During prenatal brain MRI, subtle deviations from expected maturational milestones may be observed, such as reduced or delayed gyrification or globally smaller brain tissue volumes^8^, even in the absence of macroscopic abnormalities that would explain tissue loss or atrophy^9–14^. However, these observations are largely qualitative and difficult to quantify reliably. This highlights a clinical gap: routine imaging focuses on detecting focal structural abnormalities and use biometry to assess basic growth metrics, while a more objective, holistic measurement of deviation from normative trajectories remain largely limited to research settings^15^. Direct modelling of neurological maturational timing would offer a biologically grounded framework to identify the risk of neurodevelopmental delay, assess peri-operative or postnatal effects, and guide early treatment directly after birth and during infancy, a period of increased brain plasticity.

There is emerging evidence regarding the possibility to quantify biological timelines, capturing inter-individual variability that is not explained by chronological age^16^. Molecular clocks, such as DNA-methylation age provide a proof-of-principle that biological ageing is measurable and meaningfully variable across individuals^17^. MRI offers the opportunity to depict core maturational processes at the macro-scale. Complex analyses can learn age-related patterns to derive a predicted brain age^18^ and the brain age gap (BAG), which is defined as the difference between the chronological and predicted age. The field has primarily focused on aging and brain age deviations in neurodegenerative disorders or as a consequence of established risk factors^19^.

An emerging area of research is developmental brain-age estimation using deep learning, where fetal and neonatal brain age can be reliably estimated using structural MRI data^20^. This is a promising clinical tool, as prematurity and CHD are associated with neurodevelopmental difficulties later in life, even when no apparent brain injury is detected on conventional brain MRI. In CHD, fetal MRI studies report altered cortical development in utero^13,21^, yet whether these findings reflect a global maturational delay, and the timing of the onset of the delay (i.e. prenatal or postnatal) remains unclear. In preterm birth, studies have shown that BAG increases with the degree of prematurity^22–24^, and has also been reported in conditions such as ventriculomegaly^22,25,26^. However, a key limitation in the literature is that most developmental brain age studies treat fetal and neonatal populations separately, with no studies examining the continuous trajectory throughout the perinatal period.

In this study, we develop a deep learning-based brain age framework covering the fetal–neonatal continuum to quantify deviations from normative maturational trajectories in preterm birth and CHD. We assess its generalisability across study cohorts from different centers and preprocessing pipelines, comparing models trained on structural image intensity maps, as well as segmentation-derived cortical label map representations and synthetically generated images. Normative trajectories from healthy controls serve as references for assessing maturation deviations. Using this framework, we reproduce and extend prior findings of delayed brain maturation in preterm-born neonates, serving as a biological positive control, and characterise brain age deviations in CHD across fetal and neonatal stages, examining associations with clinical severity and between preoperative and postoperative states following cardiac surgery. Finally, we link these deviations to underlying neuroanatomical patterns through spatial analyses, relating BAG differences to regional structural variation.

## Methods

### MRI datasets and image processing

Our study used retrospectively collected fetal and neonatal structural T2-weighted (T2w) brain MRI data from six datasets and three centers, spanning 1056 subjects: 440 fetuses aged 20.9-38.7 weeks gestational age (GA) and 616 neonates aged 37.0-44.9 weeks corrected gestational age (CGA). In this study, we define neonates as individuals up to 45 weeks CGA. The data comprises of neurotypical (control) fetuses and neonates, preterm-born neonates, and fetuses and neonates with CHD.

Neurotypicality was determined by (1) normality of radiological and sonography reports for the fetal datasets (Zu - rich and Shanghai fetal data), (2) normal neurodevelopment in child development evaluations at 1 or 2 years of age (Zu rich neurotypical neonates), (3) study-specific inclusion criteria in a prospective study, as detailed in the Developing Human Connectome Project (dHCP) protocol. An overview of basic data characteristics, imaging and preprocessing details are summarised in Table 1. A detailed diagram of how these datasets were assigned to various training and test sets and deep-learning models in our work is given in Figure 1.

**Table 1.** Summary of the imaging and preprocessing characteristics of each dataset for either training or testing, after quality control.

|  | Dataset size* | Age at scan (GA/CGA weeks*): mean ± SD (range) | Scanner type | T2w sequence | Resolution (mm<sup>3</sup>) | Image processing |
| --- | --- | --- | --- | --- | --- | --- |
| Neurotypical fetuses (FeTA2021) | 45 | 28.2 ± 3.9 (20.9 – 34.8) | GE 1.5T/3T (Signa Discovery MR450 and MR750) | SSTSE | 0.5 | SR reconstruction (mialSR, Simple IRTK), maternal tissue excluded |
| Control and CHD fetuses (Shanghai) | 141 | 28.8 ± 3.8 (21.0 – 38.7) | Philips 1.5T (Achieva) | SSTSE | 0.5 | SR reconstructions using NesVor |
| Control fetuses (dHCP) | 254 | 29.2 ± 3.9 (21.1 – 38.3) | Philips 3T (Achieva) | SSTSE | 0.5 | Motion correction, inhomogeneity correction, SR reconstruction (SVRTK) |
| Control and CHD neonates (Zurich) | 149 | 40.6 ± 1.7 (37.6 – 44.4) | GE 1.5T/3T (Signa Discovery MR750) | multi-slice TSE | 0.4 | Inhomogeneity correction, SR reconstructions (SVRTK) |
| Control and preterm neonates (dHCP) | 467 | 41.1 ± 1.8 (37.0 – 44.9) | Philips 3T (Achieva) | 2D TSE (ax/cor/sag) | 0.5 | Motion correction, inhomogeneity correction, SR reconstruction (SVRTK) |

\* gestational age for fetuses or corrected gestational age for neonates.
SSTSE, single-shot turbo spin-echo sequences; TSE, turbo spin-echo sequences

**Figure 1.**
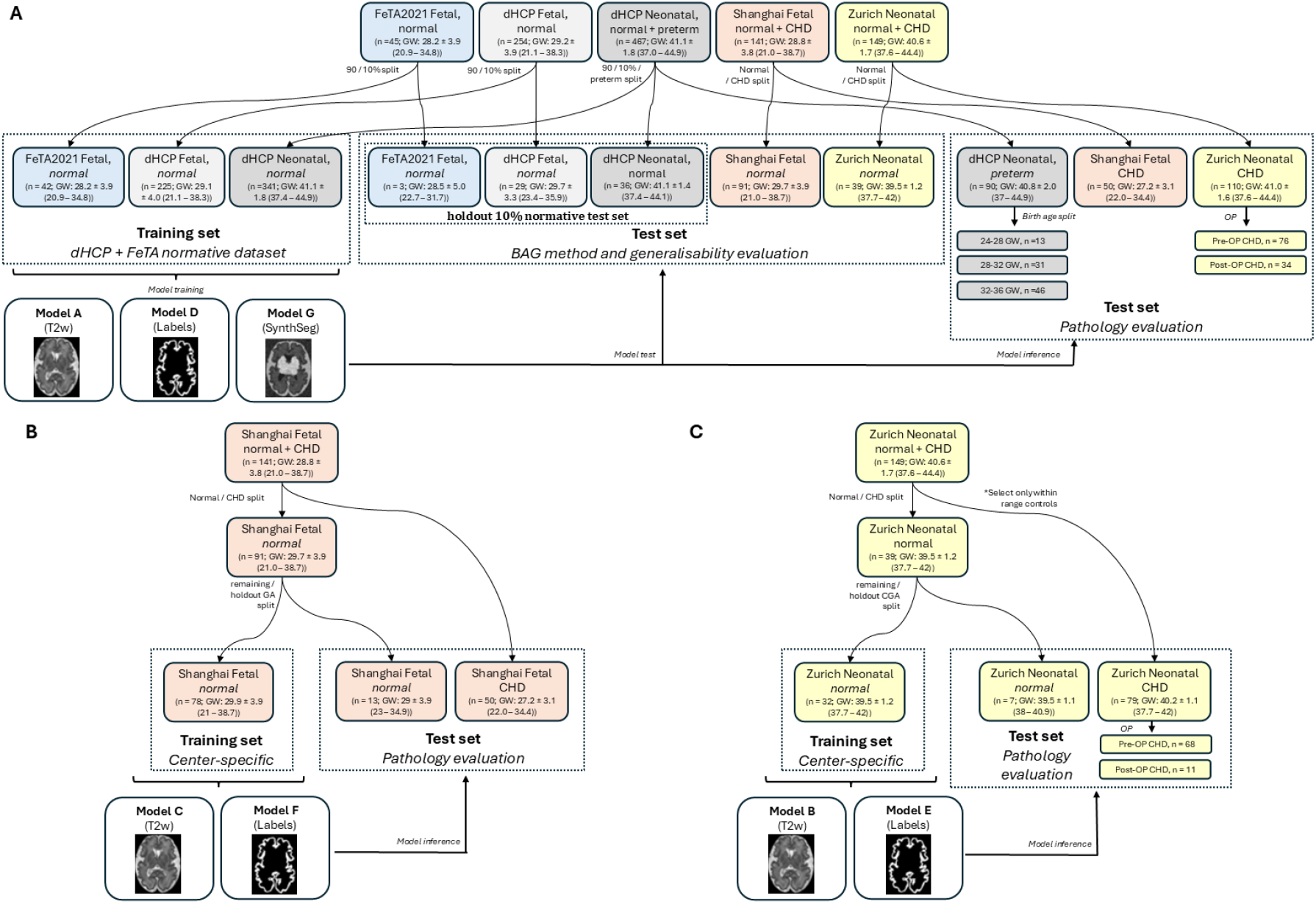
An overview of the datasets, dataset allocation, and model training strategy across multi-centric and center-specific settings. Descriptions of all trained models are provided in Table 3. **(A)** Overview of dataset allocation for models trained across the fetal–neonatal continuum. Normative fetal and neonatal MRI data from the dHCP and FeTA datasets are combined and split into training (90%) and test (10%) sets to learn a continuous developmental trajectory spanning approximately 21 to 45 weeks gestational age. Independent datasets from Shanghai (fetal) and Zurich (neonatal) serve as external control and pathological cohorts to assess cross-center generalisability and pathology evaluation, including preterm neonates and CHD. **(B)** Dataset allocation for models trained exclusively on Shanghai fetal control data to minimise site-specific bias. Control subjects are divided into training and held-out test sets using an age-stratified split. CHD fetal cases from the same center (Shanghai) are used as an independent pathology test set. **(C)** Dataset allocation for models trained on Zurich neonatal control data using an age-stratified training and test split. Neonatal CHD cases, including preoperative and postoperative scans, are evaluated as independent pathology test sets. (GW, Gestational weeks for fetuses or corrected gestational weeks for neonates; CHD, congenial heart disease; T2w, T2-weighted MRI).

### Fetal Tissue Annotation and Segmentation Challenge (FeTA) 2021 Dataset

The FeTA2021 dataset (MICCAI 2021) includes 120 fetal brain MRI scans (80 training + 40 test cases in the challenge) of healthy and pathological fetuses, acquired at the University Children’s Hospital Zurich. Each case was accompanied by manually annotated seven-class tissue segmentations, including cortical grey matter, deep grey matter, white matter, ventricles, external CSF, brainstem, and cerebellum. For the present study, 68 neurotypical cases, as defined based on the radiological report, were included. Further details regarding the dataset structure and annotations are provided in the original publications^26,27^.

### The Developing Human Connectome Project (dHCP) Fetal Dataset

In the dHCP, MRI was performed at the Evelina Newborn Imaging Center, St Thomas’ Hospital, London, UK, and includes both fetal (dHCP fetal) and term-or preterm-born neonatal (dHCP neonates) cohorts.

The dHCP fetal dataset comprises 297 T2w MRI scans acquired at 3T (21–38 weeks GA), SR reconstructed into isotropic volumes using slice-to-volume registration. Anatomical segmentations (9 tissue classes, 87 regions) were generated using the dHCP pipeline. After quality control, 265 cases with reliable segmentations were included, and labels were mapped to seven tissue classes for consistency. Minor segmentation artifacts, typically small, isolated clusters, were removed by reassigning connected components smaller than 20 voxels to their surrounding tissue. Further information on the fetal dHCP acquisition and image processing pipeline is found in^28^.

### Shanghai Fetal Control and CHD Dataset

Fetal brain MRI data from 101 controls and 56 CHD cases were acquired at the Shanghai Children’s Medical Center, parameters described in a previous publication^29^. SR produced isotropic volumes at 0.5 mm resolution using the NEsVor SR algorithm^30^. Automated segmentations were generated using BOUNTI^31^ and were mapped to seven tissue classes for consistency.

### dHCP Neonatal Dataset

The dHCP neonatal dataset includes 558 T2w scans (24–44 weeks CGA), comprising 378 term-born (born 37-44 weeks GA) and 180 preterm-born neonates (born 24-36 weeks GA). Imaging was performed on a 3T Philips Achieva scanner, acquisition details are found in^32,33^. Images were reconstructed using slice-to-volume registration (SVRTK)^34^ and segmented into 9 tissue classes and 87 regions using the dHCP neonatal pipeline^32,33^. Labels were mapped to seven tissue classes for consistency.

### Zurich Neonatal Control and CHD dataset

Neonatal MRI were obtained from the Heart and Brain study^35^ conducted at the University Children’s Hospital Zurich. This cohort includes neonates with complex CHD, with MRI acquired pre- and post-operatively, as well as healthy term-born controls scanned at term-equivalent age. The MRI dataset used for analysis, included of 40 controls, 77 CHD preoperative scans, and 36 CHD postoperative scans, acquired on a 3.0 Tesla GE MRI, further details are found in^35^. Images were subsequently SR^36^ to produce isotropic 3D volumes with a spatial resolution of 0.4 mm, followed by bias-field correction (N4, ANTs^37^) and segmentation into seven tissue classes using a custom U-Net–based pipeline with subsequent quality control and further refinement.

### CHD disease severity classification

Fetal and neonatal cardiac diagnoses encompassed a spectrum of biventricular and univentricular pathologies, including dextro-transposition of the great arteries, hypoplastic left heart syndrome, and complex aortic arch anomalies. Cases were classified according to Clancy severity categories (I–IV) based on diagnostic descriptions of the cardiac defects, by an experienced paediatric cardiologist at the University Children’s Hospital Zurich (W.K., 20 years of experience).

The distribution of cases across Clancy categories in the Shanghai fetal and Zurich neonatal cohorts is summarised in Table 2. Neonates with CHD in the Zu rich cohort underwent lesion-specific cardiac surgery according to standardised protocols, including biventricular repair or staged univentricular palliation. All procedures were performed with cardiopulmonary bypass, with selective cerebral perfusion and moderate hypothermia when indicated. Postoperative care was provided in a specialised cardiac paediatric intensive care unit (ICU), and all neonates were clinically stable at the time of MRI. Further information on the cohort is found in previous publications^35^.

**Table 2.** Overview of the models trained on neurotypical data for brain age estimation. “n” denotes the number of subjects (fetuses or neonates).

| Clancy | Shanghai fetuses (n) | Zurich neonates (n) |
| --- | --- | --- |
| I | 20 | 51 |
| II | 26 | 10 |
| III | 4 | 4 |
| IV | 0 | 11 |

### Ethical statement

Informed written consent was obtained from all parents or caregivers. Ethical approval was granted by the Canton of Zurich (2016–01019, 2022–0115; Heart and Brain study: KEK StV-23/619/04) in accordance with the Declaration of Helsinki and Good Clinical Practice guidelines. The dHCP study was approved by the United Kingdom National Research Ethics Committee, and data from the Shanghai Children’s Medical Center were used under approved data-sharing agreements (BASEC 2019-01993; Shanghai IRB).

### Image preprocessing steps for brain age estimation

T2w brain images were skull-stripped using corresponding segmentation labels and linearly registered to a 35-week postnatal T2w template from the King’s College London (KCL) neonatal atlas^38^ using affine transformation (FLIRT, 12 degrees of freedom) to account for global differences in orientation and scale. Cases with registration failure (∼2%) were excluded. After quality control, final dataset composition and training–test splits are summarised in Figure 1. Images and labels were cropped to a common bounding box (99 × 136 × 117 voxels) to remove non-informative background (Figure 2A). An overview of the image processing steps is shown in Figure 2A.

**Figure 2.**
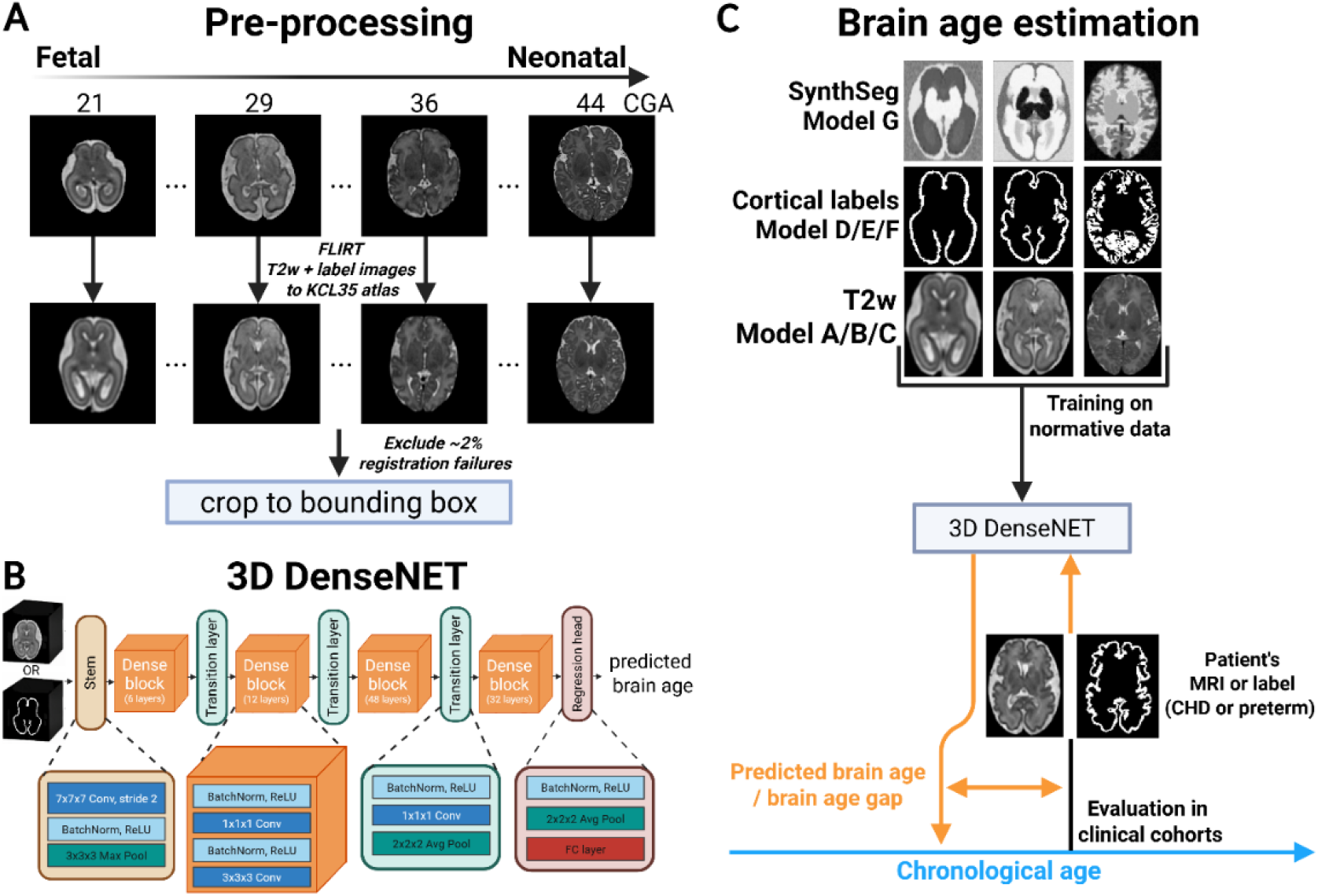
**(A) Image postprocessing steps.** Representative T2w MRI scans showing fetal to neonatal development are registered to a common reference space using affine registration to the 35^th^ week neonatal brain KCL atlas (KCL35), aligning both T2w MRIs and corresponding segmentation-derived label images. All images are subsequently cropped to a fixed bounding box to remove non-informative. **(B) DenseNET architecture**. A 3D DenseNet201 architecture implemented in the MONAI framework is used to predict brain age from volumetric MRI data. The network begins with a 7 × 7 × 7 convolutional stem with stride 2, followed by batch normalisation, rectified linear unit activation, and a 3 × 3 × 3 max pooling layer for initial feature extraction and downsampling. This is followed by four dense blocks containing 6, 12, 48, and 32 bottleneck layers, respectively. Each bottleneck layer consists of batch normalisation and rectified linear unit activation, followed by a 1 × 1 × 1 convolution and a 3 × 3 × 3 convolution, with dense connectivity ensuring that each layer receives the concatenated feature maps from all preceding layers. Transition layers between dense blocks perform feature compression using 1 × 1 × 1 convolutions and spatial downsampling via 2 × 2 × 2 average pooling. A final regression head, comprising batch normalisation, rectified linear unit activation, global average pooling, and a fully connected layer, maps the extracted features to a single continuous prediction of brain age. **(C) Brain age estimation overview**. Models are trained on normative datasets using three input representations: T2w images, cortical segmentation-based label images, and synthetically generated images. The trained networks are applied to clinical MRI data from preterm and CHD cohorts. Predicted brain age is compared with chronological age to compute the brain age gap (BAG), defined as predicted minus chronological age, which serves as a marker of maturational deviation.

### Training

We used a 3D DenseNet201 architecture (Figure 2B) implemented in MONAI to perform brain age estimation (Figure 2C). Seven distinct models were trained (Table 3), with dataset allocation and training strategy illustrated in Figure 1. The first model, designed for brain age estimation across the fetal-neonatal continuum (Figure 1A), used normative T2w data covering 20.9 – 44.9 weeks CGA; n = 676, comprising 377 term-born dHCP neonates, 254 dHCP fetuses, and 45 neurotypical FeTA fetuses. This dataset was further divided into a 90/10 training–test split (n = 608/68).

**Table 3.** Overview of the models trained on neurotypical data for brain age estimation. “n” denotes the number of subjects (fetuses or neonates). Gestational age (GA) and corrected gestational age (CGA) are reported in weeks as ranges.

|  | Dataset size | Representation | Cohort | Fetuses, n (GA weeks, range) | Neonates, n (CGA weeks, range) |
| --- | --- | --- | --- | --- | --- |
| **Model A** | 608 | T2w | dHCP fetuses / FeTA2021 fetuses / dHCP neonates | 225 / 42 (20.9 – 44.9) / (20.9 – 34.8) | 341 (37.4 – 44.9) |
| **Model B** | 32 | T2w | Zurich control neonates | - | 32 (37.7 – 42) |
| **Model C** | 78 | T2w | Shanghai control fetuses | 78 (21 – 38.7) | - |
| **Model D** | 608 | Cortical labels | dHCP fetuses / FeTA2021 fetuses / dHCP neonates | 225 / 42 (20.9 – 44.9) / (20.9 – 34.8) | 341 (37.4 – 44.9) |
| **Model E** | 32 | Cortical labels | Zurich control neonates | - | 32 (37.7 – 42) |
| **Model F** | 78 | Cortical labels | Shanghai control fetuses | 225 / 42 (20.9 – 44.9) / (20.9 – 34.8) | 341 (37.4 – 44.9) |
| **Model G** | 608 | SynthSeg | dHCP fetuses / FeTA2021 fetuses / dHCP neonates | 225 / 42 (20.9 – 44.9) / (20.9 – 34.8) | 341 (37.4 – 44.9) |

Given the challenges of generalisability across datasets from different institutions, two additional models were trained on center-specific T2w datasets (Figure 1B,C), comprising the Shanghai fetal control cohort (n = 91, 21.0 – 38.7 weeks GA), and the Zurich neonatal control cohort (n = 39, 37.7 – 42 weeks CGA).

Given the limited cohort sizes, an age-stratified subsampling was used to derive representative held-out test sets spanning the developmental range covered by the normative training data. For the Shanghai fetal cohort, subjects closest to each GA between 23 and 35 weeks were selected, resulting in a test set of 13 cases. For the Zurich neonatal cohort, neonates closest to each half week between 38 and 41 weeks corrected gestational age were selected, yielding a test set of 7 cases. The remaining subjects constituted the corresponding center specific training sets, comprising 79 Shanghai fetuses (21 – 38.7 weeks GA) and 32 Zurich neonates (37.7 – 42 weeks CGA).

To assess the impact of image representation on generalisability, we additionally trained networks using segmentation-derived label maps rather than T2w images, which are more susceptible to institution-specific variations in image intensities arising from differences in scanners, acquisition protocols, and preprocessing pipelines. We used the cortical label segmentations, which may capture developmental changes in cortical folding. Accordingly, three additional networks were trained analogous to Models A–C, but using cortical label segmentation inputs: Model D (normative fetal–neonatal cortical label model), Model E (Shanghai fetal control cortical label model), and Model F (Zurich neonatal control cortical label model).

Finally, a seventh model (Model G) was trained on synthetic T2w images generated using the SynthSeg framework^39,40^, with randomised contrast and resolution. Synthetic images were derived from the same seven-class segmentation label maps used for the normative dataset (dHCP and FeTA). To avoid introducing age-related morphological bias, we excluded scaling, shearing, rigid transformations, non-linear deformations, and bias fields^41^.

All networks were trained using the Adam optimiser^42^ with Smooth L1 loss, defined as the loss between the predicted brain age ŷ and the ground truth age *y* was computed as:

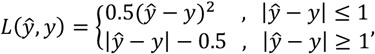

where *ŷ* is the predicted brain age and *y* the ground truth agedesigned for brain age estimation across the fetal-neonatal. Model G was trained with a learning rate of 1 × 10^−4^ and batch size 8 for 5,000 epochs, corresponding to approximately 3.04 million distinct training samples (5,000 × 608). All other models used an initial learning rate of 1 × 10^−3^ with stepwise reduction (factor 0.5, patience 10, minimum 1 × 10^−6^) and were trained for 300 epochs with batch size 8.

### Evaluation

Models trained on T2w images (Models A-C) or synthetic images (Model G) were evaluated on the corresponding T2w test sets, while label-based models (Models D–F) were evaluated exclusively on corresponding cortical label test sets. BAG was calculated as the difference between the predicted brain age and the chronological (scan) age, and mean absolute error (MAE) as the absolute value of this difference, averaged across subjects. Group level distributions of BAGs were used to characterise population level shifts into a negative direction (delay) or positive (acceleration).

### Multi-centric generalisability assessment

Generalisability across datasets and centers was first assessed by testing Model A to external control cohorts (Zurich neonates, Shanghai fetuses). For a model to be considered robust to center variability, the mean BAG in these external control cohorts was expected to be close to zero. This criterion reflects minimal systematic bias attributable to scanner differences, acquisition protocols, or preprocessing pipelines.

### Evaluation of brain age gap in preterm neonates

In preterm neonates, BAG was evaluated at term-equivalent age using dHCP data. The data was stratified by GA at birth into 3 subsets (24–28 weeks, 28–32 weeks, and 32–36 weeks; Figure 1A), to show the effect of increasing prematurity on BAG, and we tested whether there is a negative BAG in prematurity by comparing to term-born neonates of the test set.

### Evaluation of brain age gap in congenital heart disease

In CHD, BAG was assessed in fetal (Shanghai) and neonatal (Zurich) cohorts. Fetal brain maturation was assessed by comparing BAGs between fetuses with CHD from the Shanghai cohort and the Shanghai fetal controls. Postnatally, BAGs were evaluated in neonates with CHD from the Zurich cohort, with separate analyses for preoperative and postoperative MRI, each compared to healthy neonatal controls.

### Interpretability of brain age estimation

Saliency analysis was used to identify the image regions, that is, neuroanatomical structures, contributing most strongly to the brain age predictions. Saliency maps were generated only for the normative model (Model A) using a gradient-based approach^43^, in which the backward gradient of the predicted brain age with respect to the input T2w image was computed on a voxel-wise basis. Subject-level saliency maps were computed for healthy controls as well as for clinical preterm and CHD cohorts, and group-averaged saliency maps were then created by first thresholding and binarizing each subject’s saliency map at the 75^th^ percentile value, then averaging these maps across each cohort after standardising them to a canonical T2w template representing the group averaged gestational age.

### Voxel-based morphometry (VBM) analysis

VBM was carried out to relate regional volumetric variation to the BAG. A study-specific T2w template was generated using ANTs multivariate template construction^37^. All T2w images were registered to this template using sequential rigid (6 DOF), affine (12 DOF), and diffeomorphic nonlinear (SyN) transformations in ANTs^44^. Log-Jacobian (log10Jac) determinant maps derived from the nonlinear deformation fields quantified local volumetric expansion or contraction relative to the affine-normalised template space. Log-Jacobian maps were spatially smoothed with a 3 mm Gaussian kernel.

Voxel-wise associations between BAG and local deformation were assessed using FSL *randomise* tool with threshold-free cluster enhancement (TFCE; 10,000 permutations^45^), including chronological age as a covariate in the general linear model. Statistical significance was set at p < 0.05, family-wise error corrected.

### Statistical analysis

Statistical significance of group level distributions of BAG was assessed within each evaluated dataset to test for deviations between clinical and control cohorts. For all crosssectional comparisons, including generalisability assessments, preterm neonates at term equivalent age, and CHD fetal and neonatal cohorts (control vs. CHD), group means were compared using a two-sided Student t-test with unequal variances (Welch’s t-test). For CHD neonates with available longitudinal imaging, within-subject differences between preoperative and postoperative BAG were evaluated using a two-sided paired Student t-test restricted to subjects with paired scans. Statistical significance was defined as p < 0.05.

## Results

### Multi center generalisability of perinatal brain age estimation models

Models trained on T2w images (Model A) and cortical segmentation-derived label images (Model D) achieved low prediction error on normative data, but showed limited generalisability across external cohorts (Figure 3A–C). On the holdout test set, Model A achieved a MAE of 0.54 ± 0.44 weeks with a mean brain age gap (BAG) close to zero (+0.0 ± 0.7 weeks), while Model D showed comparable performance (MAE: 0.47 ± 0.34 weeks; BAG: −0.17 ± 0.56 weeks).

**Figure 3:**
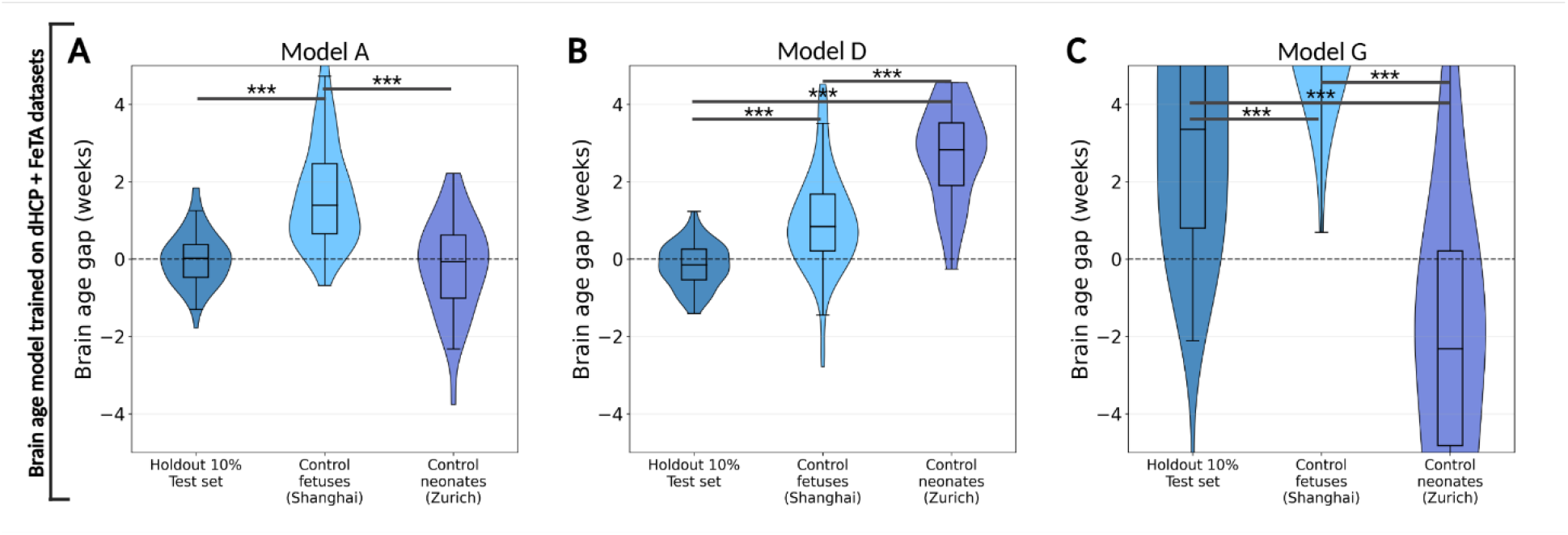
Brain age gap estimation in normally developing subjects: assessment of model generalisability across out-of-domain cohorts. Violin plots show distributions of BAG for **(A)** the T2-weighted model (Model A), **(B)** the cortical label model (Model D), and **(C)** the synthetic image-based model (Model G). For each model, performance is shown for the holdout test set (dHCP and FeTA) and independent control cohorts from Shanghai (fetal) and Zurich (neonatal). Models A and D show BAG values close to zero on the holdout test set, but show systematic positive bias in external fetal cohorts. In contrast, Model G shows inferior performance across all datasets, with large errors and systematic bias. Axes are shown on the same scale to facilitate comparison across panels. Asterisks denote statistical significance (*P < 0.05, **P < 0.01, ***P < 0.001).

In contrast, when applied to out-of-domain control cohorts, both Model A and Model D showed systematic biases, that is, a shift towards overestimating the brain age (positive BAG). For Model A, Shanghai control fetuses showed significantly elevated BAG of +1.7 ± 1.3 weeks (p = 1.27 × 10^-18^), whereas Zurich neonatal controls showed BAGs values close to zero (-0.18 ± 1,2 weeks; p = 0.38), albeit increased variance relative to the holdout dHCP test set. Model D showed similar systematic positive bias for the out-of-domain Shanghai fetal controls (+0.97 ± 1.2 weeks; p = 6.99 × 10^-12^), but in contrast to Model A, showed a marked overestimation in Zurich neonatal controls (+2.7 ± 1.25 weeks; p = 1.19 × 10^-17^). Model G showed substantial inferior performance with high errors on the holdout 10% normative test set (MAE: 3.92 ± 2.70 weeks; BAG: 3.19 ± 3.55 weeks), and very large systematic biases for both external cohorts, with BAG of 7.78 ± 2.77 weeks (p = 8.67 × 10^-15^) in the Shanghai control fetuses and -1.27 ± 3.27 weeks (p = 1.97 × 10^-10^) in the Zurich control neonates.

### Brain age estimation in preterm neonates

Preterm-born neonates imaged at term-equivalent age showed progressively more negative brain age gap (BAG) with increasing degree of prematurity, indicating delayed maturation (Figure 4). This pattern was seen consistently across both the T2w model (Model A; Figure 4A, B) and the cortical label model (Model D; Figure 4E,F). The synthetic image-based model (Model G) was not evaluated further due to its poor predictive performance (Figure 3C). All p-values refer to comparisons with term-born neonates from the test set.

**Figure 4:**
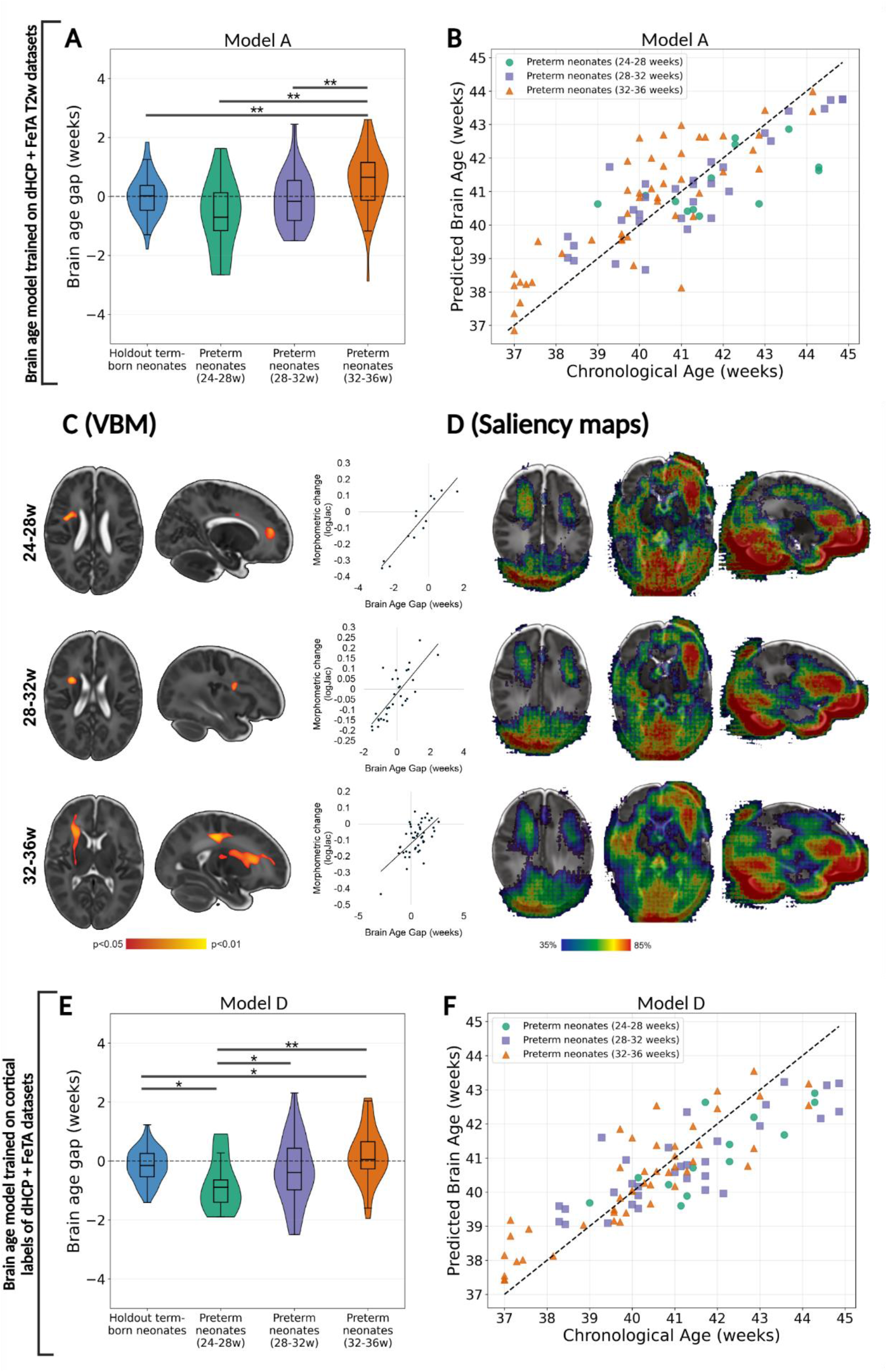
Increasing prematurity is associated with progressively more negative BAG, indicating delayed maturation: evidence from T2w and cortical segmentation-based brain age estimation models. **(A)** Group-level BAGs using T2w model (Model A). **(B)** Individual predictions versus chronological age (Model A). **(C)** VBM associations with BAG, showing regions where lower BAG correlates with reduced regional volume. Effects are localised to deep frontal white matter, internal capsule, and peri-Rolandic regions. **(D)** Saliency maps, highlighting regions contributing most strongly to age prediction. Key regions include deep white matter, brainstem, cerebellum, and CSF spaces. **(E)** Group-level BAGs using cortical label model (Model D). **(F)** Individual predictions using cortical label model (Model D). Asterisks denote statistical significance (*P < 0.05, **P < 0.01, ***P < 0.001).

Overall, both models demonstrated more negative BAG values in preterm groups relative to term-born controls. Neonates born at 24 to 28 weeks showed the largest deviations, with significantly more negative BAG for Model D (-0.79 ± 0.9 weeks; p = 0.03) compared to term-born neonates (BAG:-0.19 ± 0.59 weeks).

A similar trend was observed for Model A (−0.66 ± 1.2 weeks) compared to term-born neonates (BAG: 0.01 ± 0.70 weeks), although this did not reach statistical significance (p = 0.09). This is followed by those born at 28 to 32 weeks with BAGs of 0.11 ± 0.91 weeks (p = 0.41) for Model A and -0.34 ± 1.13 weeks (p = 0.0004) for Model D. In the 32 to 36 weeks group, Model A showed modestly positive BAG (+0.52 ± 1.03 weeks; p = 0.024), whereas Model D showed values close to zero (0.18 ± 0.91 weeks), although still significantly higher than term-born controls (p = 0.001), with a small mean difference (0.39 weeks). Scatter plots of predicted versus chronological age demonstrated a consistent downward shift in preterm groups, with a slight age-dependent effect whereby neonates scanned at slightly older ages showed more negative BAG (Figure 4B, F).

VBM analysis showed that lower BAG was associated with regional contraction (negative log-Jacobian values, TFCE-corrected p < 0.05; Figure 4C), with spatial patterns varying by degree of prematurity. In the most premature groups (24-28 and 28-32 weeks), the association was localised to frontal white matter inferior to the precentral gyrus. In the moderately preterm group (32–36 weeks), the effect extended ipsilaterally along the internal and external capsules and into deep frontal white matter beneath the preand postcentral gyri.

Saliency analysis of the models (Figure 4D) showed largely consistent attention patterns across prematurity groups, comprising CSF spaces, brainstem, cerebellum and surrounding cisterns, bilateral frontal white matter including deep precentral regions, prefrontal white matter, and occipital lobe (at both grey and white matter).

### Brain age estimation in congenital heart disease

First, we evaluated BAG in the CHD cohorts using Model A (Figure 5A-C) and Model D (Figure 5J-L), and as a next step, used the network trained specifically on only control subjects sampled from the same study center; Models B (Shanghai) and Model C (Zurich) for T2w images (Figure 5D-G); and Models E (Shanghai) and Model F (Zurich) for cortical segmentation-derived label images (Figure 5M-P).

**Figure 5:**
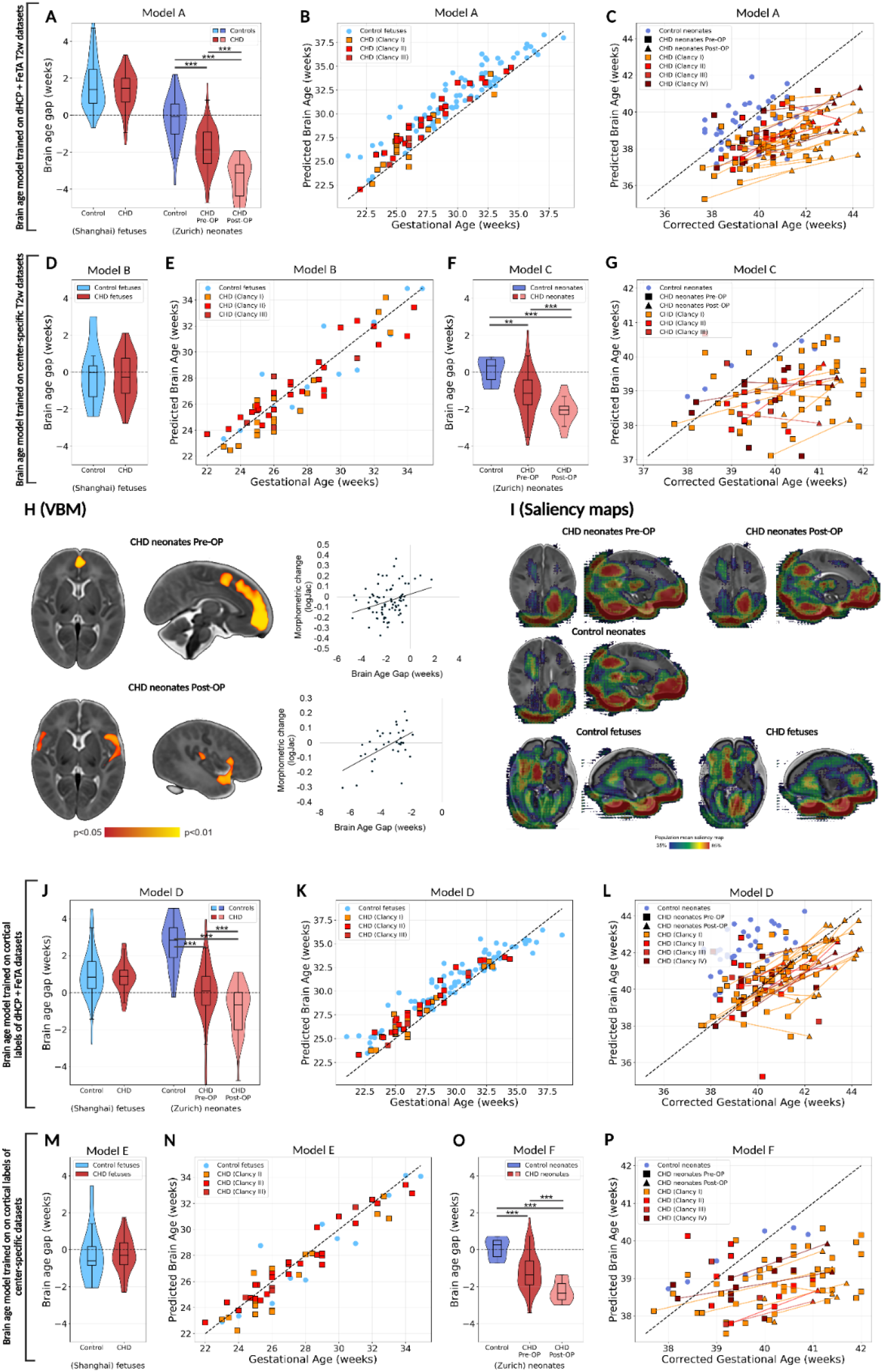
Brain age gap analysis in fetuses and neonates with congenital heart defects. Our findings reveal no delay in fetal development, but increasing BAG in pre-operative and more pronounced in postoperative MRI. **(A)** Group-level BAGs using T2w model (Model A). **(B-C)** Individual predictions versus chronological age in fetuses (B) and neonates (C), stratified by Clancy severity (I-IV) (Model A). **(D–E)**. Center-specific T2w model using Shanghai fetuses (Models B), presenting Group-level BAGs (D) and Individual predictions versus chronological age (E). **(F–G)** Center-specific T2w models using Zurich neonates (Models C), presenting Group-level BAGs (F) and Individual predictions versus chronological age (G). **(H)** VBM associations with BAG, showing regions where lower BAG correlates with reduced regional volume. Preoperative effects localise to frontal and peri-Rolandic regions, while postoperative effects shift to bilateral temporal and insular regions. **(I)** Saliency maps across fetal and neonatal CHD groups, highlighting deep white matter, thalamus, and CSF spaces as key contributors to prediction. **(J)** Group-level BAGs using cortical label model (Model D). **(K-L)** Individual predictions versus chronological age in fetuses (B) and neonates (C), satisfied by Clancy severity (I-IV) (Model D). **(M–N)** Center-specific cortical label model using Shanghai fetuses (Models E), presenting Group-level BAGs (M) and Individual predictions versus chronological age (N). **(O–P)** Center-specific T2w models using Zurich neonates (Models F), presenting Group-level BAGs (O) and Individual predictions versus chronological age (P). Asterisks denote statistical significance (*P < 0.05, **P < 0.01, ***P < 0.001).

Overall, no significant BAG shift was observed in fetuses with CHD, whereas neonates with CHD showed significantly negative BAG values compared to their respective control groups. Fetuses with CHD showed only moderately positive BAGs (+1.33 ± 1.00 weeks) comparable in magnitude to the corresponding fetal controls sampled from the same center (Shanghai data, +1.68 ± 1.34 weeks; p = 0.08) using Model A.

No differences in BAG were observed across Clancy severity categories. After birth, we found consistently significant negative BAGs in neonates with CHD compared to controls. Prior to cardiac surgery (preoperative MRI), neonates with CHD showed significantly more negative BAGs (-1.81 ± 1.18 weeks; p = 2.93 × 10^-9^) relative to healthy controls (-0.19 ± 1.24 weeks) using Model A. Similar significant differences were observed for Model D, where the controls have a high positive shift (+2.69 ± 1.26 weeks) and CHD neonates have significantly lower BAGs (+0.06 ± 1.51 weeks; p = 5.21 × 10-16).

This delay was present across all Clancy severity categories, with no clear observed stratification by lesion complexity. For Model A, a trend toward increasing BAG with advancing CGA was observed in CHD neonates (Figure 5C), whereas this effect was less apparent for Model D (Figure 5L).

Following cardiac surgery, BAG values became significantly more negative in both models (Model A: -3.56 ± 1.14 weeks, p = 1.91 × 10^-14^; Model D: -0.89 ± 1.30 weeks, p = 7.53 × 10^-5^) compared to the preoperative scans, with postoperative scans showing significantly the most negative BAG relative to controls (Model A: p = 8.40 × 10^-19^; Model D: p = 2.38 × 10^-18^). Within-subject trajectories (pre-to postoperative change of BAG in the same neonate) show an increasing negative BAG for most individuals.

When center-specific models (Models B, C, E, F) were applied to the corresponding fetal and neonatal CHD datasets, relative group differences between CHD and controls were consistent with those observed using models trained on the combined dHCP and FeTA datasets (Models A and D). This pattern was consistent for both T2w models (Model B and C; Figures 5D–G) and cortical label models (Model E and F; Figures 5M–P). These center-specific models demonstrated mean BAG values close to zero in control cohorts, albeit with higher MAEs. In Shanghai control fetuses test set, Model B yielded a BAG of −0.03 ± 1.64 weeks (MAE 1.19 ± 1.07), and in Zurich control neonates test set, Model C yielded +0.12 ± 0.70 weeks (MAE 0.62 ± 0.30). Within this unbiased reference, fetal CHD cases demonstrated BAGs also close to zero (-0.24 ± 1.21) that were not significantly different from the fetal controls (p = 0.67), whereas neonatal CHD cases showed significantly negative BAGs preoperatively (-1.28 ± 1.28; p = 0.002) compared to controls (0.12 ± 0.71) with further significant reductions following surgery (-3.13 ± 1.08; p = 2.27 × 10^-5^).

In the fetal cohorts (controls and CHD), VBM analysis found no significant association between BAG and regional morphology (TFCE-corrected p > 0.05). Neonatal CHD cases from Zürich demonstrated distinct morphometric patterns depending on perioperative status. Preoperatively, lower BAG was associated with regional contraction (negative log-Jacobian values, TFCE-corrected p < 0.05; Figure 5H), predominantly along the midline frontal regions surrounding the precentral gyri and within adjacent frontal white matter. Postoperatively, the spatial pattern shifted, with BAG-related contraction observed bilaterally in the temporal poles, insular regions, and inferior frontal lobes.

Saliency analysis of the brain age models (Figure 5I) showed consistent attention patterns across fetal and neonatal control and CHD groups: inferior CSF spaces surrounding the cerebellum, deep bilateral frontal and parietal white matter, thalamus, and inferior frontal regions. In fetal cohorts, additional attention was seen in the thalamus and occipital horns of the lateral ventricles.

## Discussion

Brain age gap provides a global measure of developmental timing. It has the potential to enable the detection of deviation from normative trajectories beyond focal structural abnormalities. In this study, we developed a deep learning-based brain age estimation framework that is applicable across the fetal–neonatal MRI continuum and tested it on two clinically relevant groups of neonates at risk for neuro-developmental impairments: preterm birth and CHD.

We found that brain age estimation across fetal and neonatal MRI is feasible using either T2w images or segmentation derived cortical labels, but remains sensitive to scanner, acquisition, and preprocessing variability, highlighting the importance of model calibration through center specific training, refinement and image representation choice. We revealed that more negative BAG values were observed in preterm-born neonates in proportion to the degree of prematurity, which may indicate a progressively greater lag in brain maturation with increasing prematurity, particularly in neonates born before 28 weeks of gestational age. In CHD, brain development appeared largely preserved in utero, whereas consistent significant delays emerge postnatally, with more pronounced negative BAG observed after cardiac surgery. No consistent link was observed between underlying cardiac severity category and BAG.

### Generalizability of perinatal brain age estimation

While showing a good performance on normally developing fetuses and neonates, our models did not generalise well across centers. Both T2w and cortical label models showed systematic biases on unseen control datasets, even when intensity-based data augmentation strategies such as gamma augmentation (not shown) were used. Models trained on SynthSeg-generated synthetic images also showed poor generalisability across datasets from different institutions. While SynthSeg has been shown feasible for age estimation in aging cohorts^41^, the random intensity contrast generation, as implemented in SynthSeg, may have been insufficient to capture developmental changes in white-grey matter contrast and evolving myelination patterns. Our findings show that robust generalisability across centers and developmental stages remains technically challenging and depends strongly on input representation. Using center-specific training data improved performance, but further refinement might be necessary in future works. Augmentation through image generation, such as by diffusion models, may improve domain robustness, but their substantial data requirements, often on the order of hundreds to thousands of training samples^46^, limits their applicability in our setting.

Our prediction accuracy (MAE of ∼0.5) is comparable with previous works in the field, where developmental brain age prediction accuracy is typically reported in the sub-week to 1–2 week range, depending on imaging modality^20,22^. BAG might be treated as a normative deviation marker rather than a diagnosis: neonatal connectome-based work has linked BAG to deviations from expected developmental trajectories^21^. Although we did not assess associations with long-term outcomes, previous work has reported that older-appearing brains relate to adverse outcomes, such as mortality^47^. At the same time, these findings highlight the importance of model calibration and outcome validation^47^.

### Brain age estimation in preterm born neonates

Consistent with prior studies^22,23,48^, we found more negative BAGs at term-equivalent age with greater degrees of prematurity, which may reflect delayed development. This aligns with MRI evidence that brain maturation can lag behind expected gestational age, suggesting earlier or ongoing injury^49^. Neonates born at 24–28 weeks showed the largest delays followed by those born at 28–32 weeks. The intensity-based model (Model A) showed slight positive values in less premature neonates, suggesting suggests that intensity-driven features, related to changing tissue contrast due to myelination patterns, may affect age prediction independently of macro-morphological maturation (e.g. gyrification). Therefore, structural MRI alone may not capture all aspects of neurodevelopmental maturation, such as micro-structural processes that evolve rapidly in the perinatal period. Such microstructural features have been shown to improve brain age estimation performance when combining structural-MRI and diffusion tensor imaging^22^. When we restricted the model to cortical shape features, the results remained reliable and showed a similar trend in preterm neonates, which could mean that cortical segmentation can be a minimalistic set of data for brain age estimation, even in case of multicentric datasets with no MRI protocol harmonization.

The negative BAG in preterm neonates is biologically plausible as a systems-level summary of dysmaturation^50^, particularly affecting white matter. Preterm brain injury is often dominated by white-matter injury and impaired myelination, in which pre-oligodendrocytes regenerate but fail to mature. This maturational arrest leads in hypomyelination and altered developmental trajectories even without overt focal lesions on MRI^51–53^.

Our analysis of the association of brain morphometry and BAG in prematurity showed a relative volume reduction in small areas in the frontal deep white matter. With decreasing GA, these reductions extended toward internal and external capsules and peri-Rolandic deep layers. This is consistent with vulnerability of early-maturing projection pathways supporting motor and cognitive control^54^. The localisation of white matter injuries may have contributed to the anatomical specificity of our findings^54,55^. The posterior limb of the internal capsule and associated corticospinal projections undergo fast myelination around term-equivalent^56,57^, providing a possible anatomical substrate for BAG related morphometric patterns in peri-Rolandic and capsular regions. However, the affected regions covered a much smaller spatial extent than the white matter injury distribution typically observed on group-level probability maps, as reported by Guo et al^58^.

The saliency maps converged on deep frontal and parietal white matter, the brainstem, cerebellum, and surrounding CSF spaces. This suggests that BAG might stem from multiple coupled processes, including white-matter dysmaturation and impaired myelination alongside coordinated volumetric growth during the late preterm period^59,60^.

### Brain age estimation in congenital heart disease

We found a negative BAG in neonates with CHD of approximately 1.5 to 2 weeks (depending on model and subject-level variability), whereas this was not observed in fetuses. This suggests a lag relative to normative developmental trajectories that becomes apparent postnatally. In utero, fetuses with CHD showed no clear stratification by disease severity or cardiac physiology, which confirmed after applying center-specific training to reduce systematic bias.

These findings suggest that, despite known alterations in cerebral growth, such as volumetric reductions^61–63^, global brain maturation as captured by brain age estimation is largely preserved during fetal life. This contrasts with some fetal MRI studies reporting subtle developmental delays in CHD, e.g. as a function of lesion subtype and cerebral oxygen delivery, rather than reflecting a uniform global pattern^64,65^. Reduced fetal cerebral oxygen delivery has been linked with smaller brain size without necessarily producing a detectable global timing shift, consistent with heterogeneous growth trajectories^13,21^. We speculate that our method became agnostic to overall brain tissue volumes due to the necessary image size standardisation steps as in other related BAG estimation works^20^, and that reported gyrification differences in CHD may be too subtle for the network to capture^67^, or gyrification patters might show too much inter-individual variability to be reliably captured by a global learning approach. However, a clear limitation is that the most severe cardiac physiologies were not included in the fetal MRI dataset (Clancy IV category) due to possible patient enrolment bias.

The slight positive BAG observed in fetal CHD may reflect absence of detectable delay rather than accelerated development. In contrast, postnatal and perioperative exposures, including cardiopulmonary bypass and perioperative instability, may impose additional physiological stress that slows maturational speed even in the absence of overt focal injury, consistent with reported perioperative brain vulnerability in CHD^68^.

Brain morphometric changes measured between pre- and postoperative MRI, which did not manifest in MRI visible cerebral lesions, have been reported to correlate with the intraoperative cerebral desaturations^69^, establishing a potential explanation for our findings. Early postoperative growth and microstructural development are particularly sensitive to the immediate ICU course, with postoperative brain growth impairments and brain injury associations reported with perioperative physiology and clinical course in CHD^70^. Although fetal CHD is linked to reduced cerebral oxygen delivery and smaller brain size, the fetal environment may permit partial compensation, or alterations may remain regionally specific and not detectable by a global metric^71^.

In CHD neonates, BAG associated morphology shifts from midline frontal and peri-Rolandic regions preoperatively to bilateral temporal and frontal regions postoperatively might suggest a transition from predominantly developmental dysmaturation to additional perioperative vulnerability^72–74^. Early alterations in frontal and projection pathways are consistent with delayed maturation of rapidly developing white matter tracts, whereas postoperative involvement of temporal and insular regions may reflect the increased susceptibility of association cortex and territories to hypoperfusion and fluctuations in cerebral oxygen delivery during and after cardiac surgery^70,75^. These regions are characterised by high metabolic demand and later developmental trajectories, making them particularly vulnerable to perioperative physiological stress^59,76^.

Our results show that the morphometric associations with BAG in CHD and preterm neonates involve overlapping white matter regions, although the underlying mechanisms likely differ. Preterm white matter injury reflects selective vulnerability of pre-oligodendrocytes and arrested maturation in rapidly developing periventricular white matter^77^, while CHD is associated with impaired cerebral oxygen delivery and delayed brain growth^71^. As BAG represents a global index of maturational timing, it may capture the cumulative effect of delayed white matter expansion and early myelination during the late fetal and early neonatal period in both conditions.

## Limitations

Although we used multi-centric data for training of the models, residual site-specific effects remained, which is a limitation of our study. As a solution, center-specific training may have eliminated systematic bias, but limits generalisability and highlights the need for larger, harmonised normative datasets spanning the fetal–neonatal continuum.

Brain age estimation based on MRI provides a global summary measure of brain maturation but may not capture brain region specific, neural connectivity or microstructural abnormalities that are known to contribute to neurodevelopmental outcomes. As shown by differences between T2w and segmentation-based models, structural MRI alone may incompletely represent maturational processes. Incorporating diffusion or functional MRI may improve sensitivity, but may further create generalisability challenges due to the more complex acquisition and processing of such MRI data. A further limitation is that our sample sizes within specific clinical cardiac physiology sub-groups were limited.

An important limitation is that normality was mostly defined using radiological reports or appearance in, without consistent long-term clinical outcome data. Subtle or later-emerging impairments cannot be excluded. Ideally, training would rely on “super normal” individuals with documented normal development at 2 year follow-up, and future models could be further surrogated by molecular markers of biological aging, which remains an ethical and practical challenge. The conceptual simplicity of BAG (lag vs. acceleration) carries limitations, too, as neurodevelopmental deviations may reflect alternative trajectories, individual catch-up, or compensatory neuroplastic mechanisms that are not fully captured by a single global metric. In addition, no systematic neurodevelopmental outcome measures were available in either control or clinical cohorts, and therefore the clinical relevance of BAG remains indirect. Therefore, BAG cannot be interpreted as a validated surrogate of cognitive or functional outcome at the individual level.

## Conclusion

Developmental brain age estimation potentially reframes perinatal MRI from lesion detection toward quantification of maturational timing. The divergence between preserved fetal maturation and postnatally emerging delay in congenital heart disease might underline the potential of fetal or early neonatal life as a clinically actionable window for neuroprotection and targeted follow up. Our findings emphasize the importance of multi-center harmonisation to allow reliable use across scanners and clinical settings: with appropriate cross-site calibration, brain age estimation might become generalisable. This way, brain age metrics could support early risk stratification and function as quantitative outcome measures in studies aiming to improve neurodevelopment.

## Acknowledgement

During preparation of the initial draft, the first author used ChatGPT (OpenAI) to improve readability. All text was subsequently reviewed and edited by the authors, who take full responsibility for the content of the publication.

## Data availability

The data that support the findings of this study are not publicly available due to ethical and data protection restrictions related to human subjects. Access may be granted upon reasonable request to the corresponding author, subject to institutional approval and the establishment of a data transfer and use agreement (DTUA). Data derived from the Developing Human Connectome Project (dHCP) are subject to the data access policies of that consortium and must be requested directly through their established application procedures.

Training and inference code, as well as the trained models necessary to reproduce the main analyses, are available in a public repository at: https://github.com/Mishakaan-dorp/Developmental-brain-age-estimation.

